# Network-Based Analysis of Fibromyalgia-Associated Genes Identifies Candidate Molecular Targets and Enriched Biological Pathways

**DOI:** 10.64898/2026.09.18.26363374

**Authors:** Subham Sarkar, Anand Srinivasan

**Affiliations:** Department of Pharmacology, All India Institute of Medical Sciences, Kalyani, India; Department of Pharmacology, All India Institute of Medical Sciences, Bhubaneswar, India

**Keywords:** Network Pharmacology, Fibromyalgia, TLR, TNF, IL6, protein-protein interaction, functional enrichment analysis, gene-disease association

## Abstract

**Background:** Fibromyalgia (FM) is a disorder characterized by chronic pain and somatic symptoms that are not fully understood. Central sensitization, a process by which the central nervous system amplifies pain signals, is a key aspect of the pathophysiology of fibromyalgia. Despite its prevalence, effective treatment options are limited and the underlying pathophysiological mechanisms are not fully understood. Network pharmacology, which analyzes the interactions between diseases, targets, and biological pathways, can be used to identify potential molecular targets and pathways associated with fibromyalgia. In this study, we aimed to identify potential hub genes and the underlying biological pathways involved in fibromyalgia.

**Methods:** Four online databases, GeneCards, DisGeNET, OMIM, and PharmGKB, were initially screened for genes associated with fibromyalgia. GeneCards and DisGeNET were subsequently used to identify common genes, which were used to generate a protein association network using the STRING database. Cytoscape software was used to analyze the network, and the CytoHubba plugin was used to identify hub genes based on degree and betweenness centrality. Functional enrichment analysis was performed using ShinyGO to identify significantly enriched KEGG pathways.

**Results:** TNF and IL6 were identified as the top two hub genes with the highest degree of centrality. Functional enrichment analysis identified the Toll-like receptor (TLR) signaling pathway as a significantly enriched pathway among the prioritized hub genes, with a fold enrichment of 6.78 and FDR of 0.0346.

**Conclusion:** This study identified TNF, IL6 and other highly connected genes, along with TLR signaling as an enriched pathway in the fibromyalgia-associated gene network. These findings provide potential molecular targets and pathways for further investigation into the pathophysiology of fibromyalgia. Further experimental and clinical studies are required to independently validate these findings and determine their therapeutic relevance.

## Introduction

Practitioners in the clinical setting frequently encounter patients experiencing pain and somatic symptoms that cannot be fully accounted for by the observed levels of damage or inflammation in peripheral tissues (1). When no discernible cause is identified, these individuals often receive a diagnostic classification implying localized bodily pain such as chronic low back pain, headache, or temporomandibular joint disorder. Alternatively, the diagnosis may indicate an underlying pathological irregularity, such as endometriosis or facet syndrome, which may or may not be directly related to the individual’s pain. In some cases, patients may be informed that no physiological abnormalities are detected and may be considered to have predominantly psychosomatic symptoms without being offered appropriate treatment (2).

Fibromyalgia (FM) is a disorder of pain regulation. It is often classified under the term central sensitization (3). Central sensitization amplifies neural signaling within the central nervous system (CNS), resulting in pain hypersensitivity (4). In patients with FM, central sensitization may be accompanied by peripheral pain mechanisms, including neuropathic pain, small fiber neuropathy (SFN) (5), and nociceptive pain (such as an inflamed joint). Fibromyalgia is estimated to affect 2– 4% of the population (6). Women are affected approximately twice as often as men. The pathophysiology of fibromyalgia has not yet been fully elucidated, and several theories have been proposed (5,6).

FM shares several clinical and pathophysiological features with other common pain disorders that are considered more central than peripheral pain conditions, such as migraine, tension headaches, temporomandibular joint disorder, vulvodynia, irritable bladder, and irritable bowel syndrome. The clinical characteristics of these conditions include widespread pain, fatigue, and sleep and mood disturbances. These conditions share common genetic and CNS pain-processing mechanisms with FM.

Network pharmacology is a discipline developed based on systems biology and polypharmacology. It focuses on the multi-target process in which drugs act rather than the traditional single-target process (8). This approach can be used to study the interactions between diseases, molecular targets, and biological pathways in the form of a network, thereby providing a broader view of disease-associated molecular mechanisms (8).

The aim of the present study is to use a network pharmacology approach with functional enrichment analysis to identify disease-associated hub genes and biological pathways involved in fibromyalgia and to identify potential targets for further investigation.

## Methodology

This is a computational biology study utilizing a network pharmacology to fulfill the objectives of the study (Figure 1).

**Figure 1:**
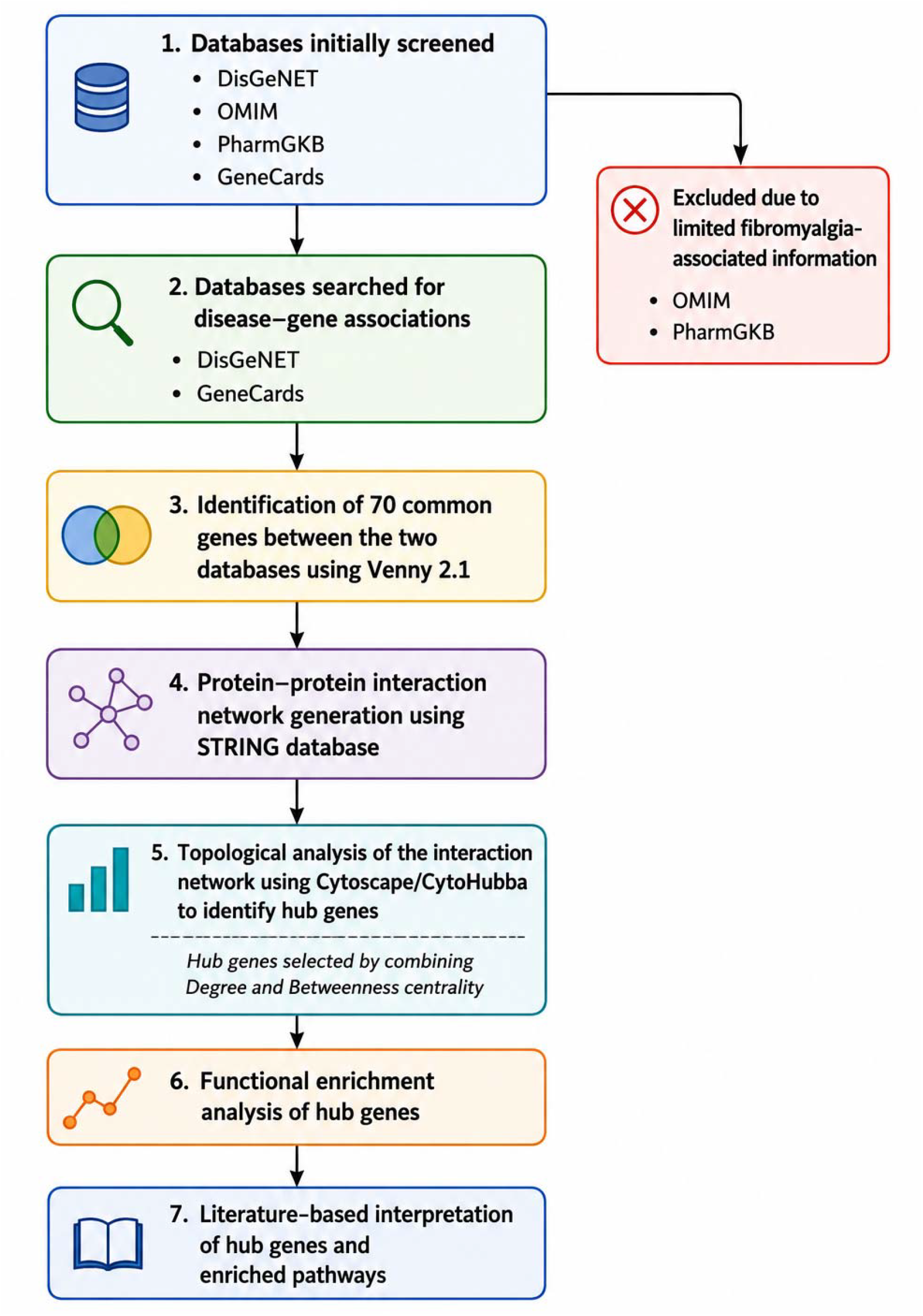
Flowchart of the study workflow. Note: DisGeNET, Disease Gene Network; OMIM, Online Mendelian Inheritance in Man; PharmGKB, Pharmacogenomics Knowledge Base.

### A. Finding disease-gene association

Disease-gene association is a type of biological data that is used to infer the connections between a disease and associated genes. This information can be obtained from various sources, including gene variants associated with diseases, biological pathways, gene expression data, biomedical ontologies, and text mining (9).

Four online databases, GeneCards version 5.17 (https://www.genecards.org/), OMIM (https://www.omim.org/), PharmGKB (https://www.clinpgx.org/), and DisGeNET version 7.0 (https://www.disgenet.org/), were initially screened for genes associated with fibromyalgia using the keyword “Fibromyalgia.” OMIM and PharmGKB were excluded from further analysis because of the limited amount of available information related to fibromyalgia in these databases. GeneCards and DisGeNET were therefore used for the subsequent analysis. GeneCards and DisGeNET retrieved 293 and 143 genes, respectively. Venny was used to identify the common genes between GeneCards and DisGeNET, resulting in 70 common genes, which were used for further analysis.

### B. Protein association network

Protein-protein interaction networks (PPIN) are mathematical models that depict how proteins physically interact within a cell. As proteins play a fundamental role in biological functions, protein-protein interactions (PPIs) are crucial for determining the molecular and cellular mechanisms that govern both healthy and diseased states in organisms (10).

To generate the fibromyalgia-associated protein network, the list of genes common to GeneCards and DisGeNET obtained from the intersection using Venny 2.1 was imported into the STRING database (https://string-db.org/). *Homo sapiens* was selected as the organism, with a minimum interaction confidence score of 0.400. The resulting network was exported in tab-separated value (TSV) format for further network analysis. Of the 70 common genes used as input, 67 were represented as nodes in the resulting STRING network.

### C. Network analysis

In protein association networks, proteins are represented as nodes, while interactions are represented as edges. The topological arrangement of the protein association network, as per graph theory, offers fundamental insights into the network properties that may provide information about the biological functions of the network (11).

The network from the STRING database was then imported to Cytoscape (12) as a tab-separated value (TSV) file. Topological analysis of the network was performed using the CytoHubba plugin in Cytoscape.

Two pre-set criteria were used for network analysis:

a. **Degree centrality**
b. **Betweenness centrality**

CytoHubba was used to assess degree and betweenness centrality, and the top 10 hub genes were prioritized by considering both measures. Genes with high degree centrality were considered important because they interact with a larger number of other genes within the network. Betweenness centrality is based on the shortest paths between nodes and identifies genes that may act as bridges connecting different parts of the network.

### D. Functional enrichment analysis

Functional enrichment analysis seeks to identify biological annotations that are over-represented in a list of genes compared to a reference background. These annotations are employed to interpret the molecular mechanisms and biological processes that are associated with the disease condition under study (13).

After obtaining the hub genes, functional enrichment analysis was performed using the ShinyGO 0.80 (14) platform with *Homo sapiens* selected as the species. The top 10 hub genes obtained from the network topological analysis were imported into the ShinyGO platform. The list of common fibromyalgia-associated genes obtained from GeneCards and DisGeNET, which was used to generate the protein association network, was used as the background gene set.

The following filtering criteria were applied in ShinyGO: (a) FDR cutoff = 0.05, (b) pathways to show = 10, (c) pathway size minimum = 10, (d) pathway size maximum = 2000, and (e) database = KEGG.

These criteria were used to focus the analysis on statistically significant pathways and to limit the inclusion of very small or very large pathways.

## Results

### A. Gene-disease association

OMIM and PharmGKB were excluded from further analysis because of the limited amount of available information on fibromyalgia disease-gene association.

GeneCards and DisGeNET were therefore used for further analysis. GeneCards and DisGeNET retrieved 293 and 143 genes, respectively. Using Venny 2.1, 70 genes common to both databases were identified for fibromyalgia (Figure 2).

**Figure 2:**
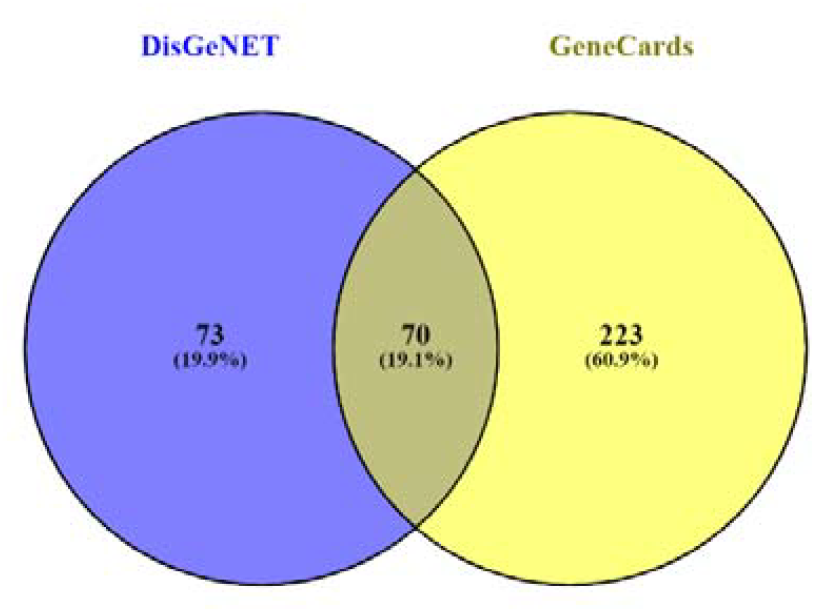
Venn diagram showing the common genes between DisGeNET and GeneCards for fibromyalgia.

### B. PPI Network

The 70 common genes identified from GeneCards and DisGeNET were imported into the STRING database to generate the protein-protein interaction network. The resulting network consisted of 67 nodes and 636 edges (Figure 3). The network had an expected number of edges of 140, with an average local clustering coefficient of

**Figure 3:**
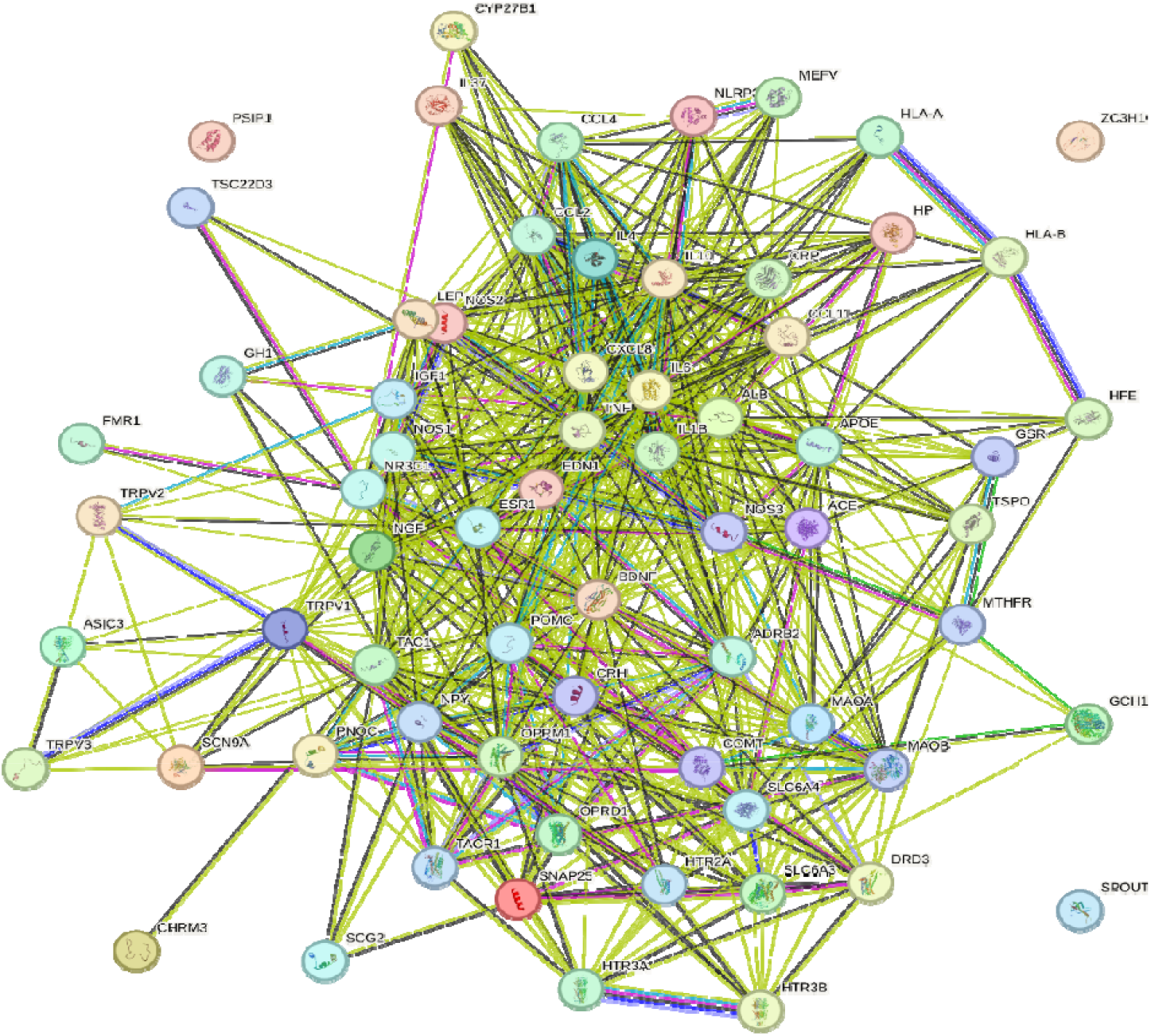
Protein association network generated using STRING from the common fibromyalgia-associated genes.

0.68 and an average node degree of 19. The PPI enrichment p-value was<1.0 × 10-16, indicating that the observed protein associations were greater than expected for a random set of proteins (Table 1).

**Table 1:** Summary statistics of network.

| S.NO | Parameter | Value |
| --- | --- | --- |
| 1 | Number of nodes | 67 |
| 2 | Number of edges | 636 |
| 3 | Expected number of edges | 140 |
| 4 | Average local clustering coefficient | 0.68 |
| 5 | Average node degree | 19 |
| 6 | PPI enrichment p-value | $<1.0 \times 10^{-16}$ |

### C. Finding hub genes

After importing the PPI network in TSV format into Cytoscape, the CytoHubba plugin was used to perform topological analysis as described in the methodology. CytoHubba was used to assess degree and betweenness centrality, and the top 10 hub genes were prioritized by considering both measures (Figure 4). The degree centrality values of the identified hub genes are presented in Table 2.

**Table 2:** Summary of hub genes ranked according to degree centrality.

| Rank | Hub gene | Degree centrality |
| --- | --- | --- |
| 1 | TNF | 45 |
| 1 | IL6 | 45 |
| 3 | IL1B | 42 |
| 4 | ALB | 41 |
| 4 | BDNF | 41 |
| 6 | TAC1 | 38 |
| 7 | IL10 | 36 |
| 8 | POMC | 35 |
| 9 | NGF | 34 |
| 10 | CXCL8 | 33 |

**Figure 4:**
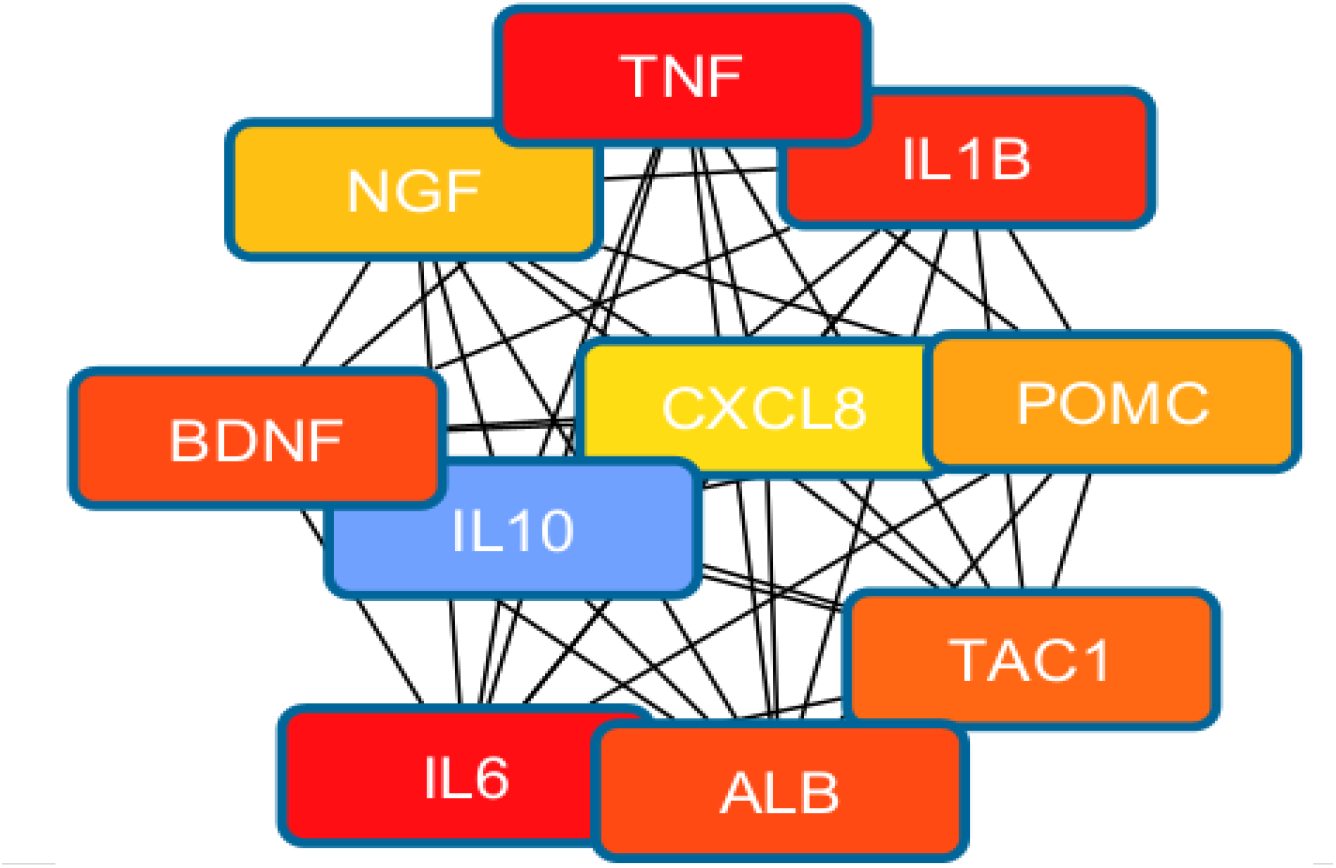
Interaction network of the top 10 hub genes identified from the fibromyalgia-associated protein network.

According to degree centrality, TNF and IL6 were the top two hub genes, with a degree of 45 for both genes, indicating the highest number of direct interactions within the network among the identified hub genes.

### D. Result of Enrichment analysis

In the ShinyGO platform, functional enrichment analysis of the top 10 hub genes identified several significantly enriched KEGG pathways. The Toll-like receptor signaling pathway was among the pathways with the highest fold enrichment, with a fold enrichment of 6.78 and FDR of 0.0346. Three of the analyzed hub genes, IL1B, IL6, and CXCL8, were associated with this pathway (Table 3).

**Table 3:** Summary of functional enrichment analysis.

| Pathway | Genes | No. of genes | Pathway genes | Fold enrichment | FDR |
| --- | --- | --- | --- | --- | --- |
| Toll-like receptor signaling pathway | IL1B, IL6, CXCL8 | 3 | 103 | 6.78 | 0.0346 |

## Discussion

At present, the pathogenesis and molecular mechanisms of fibromyalgia have not been fully elucidated. However, from the research progress made thus far, it is known that fibromyalgia (FM) is a disorder of pain regulation and is associated with central sensitization (3) (15). Evidence suggests that one of the key mechanisms behind the induction & maintenance of central sensitization is neuroinflammation in the central & peripheral nervous system(16).Neuroinflammation is identified by vascular changes that increase permeability, activation of glial cells, infiltration and activation of leukocytes, and increased production of inflammatory mediators such as cytokines and chemokines.(17) Evidence indicates that cytokines, chemokines, and other mediators produced by glial cells can initiate or sustain central sensitization, which contributes to the emergence of widespread pain(16). Neuroinflammation seems to arise from a two-way communication between nociceptors and various non-neuronal cells, particularly immune and glial cells. In this process, cytokines and inflammatory mediators from immune and glial cells activate and sensitize nociceptors. At the same time, nociceptors can also release cytokines and chemokines, which play a crucial role in immune modulation(16).

Several observational and biological studies suggest that chronic widespread pain and fibromyalgia (FM) may have a genetic basis (18). First-degree relatives of patients with FM are 8.5 times more likely to have FM than relatives of patients with rheumatoid arthritis (19). A subsequent genome-wide profiling found that, compared with controls, patients with FM had differences in the expression of 421 genes, many of which were involved in pain processing (20).

In this study, we used a network pharmacology approach to identify potential targets and biological processes involved in the pathophysiology of fibromyalgia. We identified hub genes by topological analysis of the protein association network generated in our study. Hub genes are highly connected genes within a biological network and may have an important position in the network(21). Therefore, highly connected genes may represent potential candidates for further investigation in disease-associated pathways and therapeutic research (22). In our study, TNF and IL6 were the top two hub genes identified, with the highest degree of centrality (45 for both). Based on their high connectivity in the fibromyalgia-associated network, we hypothesize that TNF and IL6 may have an important role in the complex molecular mechanisms associated with fibromyalgia.

Previous meta-analytic evidence provides support for the involvement of inflammatory cytokines in fibromyalgia. One meta-analysis observed that patients with fibromyalgia had higher plasma IL-6 levels compared with controls (standardized mean difference [SMD (95% CI)] = −0.34 [−0.64 to −0.03]; p = 0.03) (23). Another meta-analysis found significant differences in peripheral blood cytokine profiles in patients with fibromyalgia compared with healthy controls, including increased TNF-α (SMD [95% CI] = 0.36 [0.12 to 0.60], p = 0.0034), IL-6 (SMD [95% CI] = 0.15 [0.003 to 0.29], p = 0.045), and IL-8 (SMD [95% CI] = 0.26 [0.05 to 0.47], p = 0.01)(24). These findings are concordant with our identification of TNF and IL6 as highly connected genes in the fibromyalgia-associated network.

Currently, several pharmacological therapies are used for the management of fibromyalgia, including tricyclic antidepressants such as amitriptyline, serotonin-norepinephrine reuptake inhibitors such as duloxetine, and gabapentinoids such as pregabalin (25)

From our network analysis, IL6, TNF, and IL10 were identified among the highly connected hub genes in the fibromyalgia-associated network. IL6 and TNF are pro-inflammatory cytokines, whereas IL10 has predominantly anti-inflammatory effects. These findings suggest that dysregulation of inflammatory signaling may contribute to the molecular mechanisms associated with fibromyalgia. Previous evidence has shown that antidepressant treatment can decrease peripheral levels of inflammatory cytokines, including IL-6, TNF-α, and IL-10, in patients with major depressive disorder (26). This provides a possible biological explanation for the effects of antidepressant drugs on inflammatory signaling, although whether such mechanisms contribute to the clinical effects of antidepressants in fibromyalgia requires further investigation.

From the fibromyalgia-associated network, we also found that BDNF was one of the highly ranked hub genes, with a degree of 41, suggesting that it may have an important position in the molecular network associated with FM.

Our finding is similar to a clinical study in which serum BDNF levels were studied in patients with chronic fatigue syndrome with comorbid fibromyalgia. Compared to controls, serum BDNF levels were higher in patients with CFS/FM (F = 15.703; mean difference = 3.31 ng/ml [95% CI 1.65 to 4.96]; P = 0.001)(27) The study also found that higher serum BDNF levels were associated with symptoms and widespread hyperalgesia. These findings suggest that altered BDNF levels may be involved in the pathophysiology of fibromyalgia.

In the functional enrichment analysis, the Toll-like receptor signaling pathway was among the significantly enriched pathways, with a fold enrichment of 6.78 and FDR of 0.0346. Previous studies have suggested a role of TLR signaling in fibromyalgia. In a pilot human study, patients with fibromyalgia showed an altered immune response following TLR4 activation compared with healthy individuals, suggesting that altered TLR4-related immune responses may be involved in fibromyalgia (28). These findings provide biological support for further investigation of TLR-related signaling in fibromyalgia.

Naltrexone, an opioid receptor antagonist, is commonly used for opioid and alcohol dependence. Previous studies have suggested that the opioid-inactive (+)-isomer naltrexone has activity at Toll-like receptor 4 (TLR4), based on evidence from biophysical, *in vitro, in silico*, and *in vivo* studies (30). In another study, naltrexone inhibited the production of inflammatory cytokines such as IL-6 and TNF-α by peripheral blood mononuclear cells (PBMCs) following stimulation with ligands for TLR7, TLR8, and TLR9, but not following stimulation with a TLR4 ligand. These findings suggest that the effects of naltrexone on inflammatory signaling may involve mechanisms beyond direct TLR4 inhibition.

There have been pilot clinical trials in which low-dose naltrexone was investigated for fibromyalgia and neuropathic pain, with reductions in symptoms reported in some studies(31). In one study, low-dose naltrexone reduced fibromyalgia symptoms in the entire cohort, with a greater than 30% reduction in symptoms compared with placebo.(32) In another study, low-dose naltrexone was associated with a significantly greater reduction in baseline pain compared with placebo (28.8% versus 18.0% reduction; P = 0.016). It was also associated with improved general satisfaction with life (P = 0.045) and mood (P = 0.039), but not fatigue or sleep (33)

## Limitations

This study has some limitations. First, the analysis was based on publicly available disease-gene association databases and computational network analysis, and therefore the identified hub genes and pathways represent associations rather than experimentally validated causal mechanisms. Second, the results are dependent on the genes and interactions available in the databases used and on the parameters applied during network construction and enrichment analysis. Third, the study did not include experimental validation of the identified hub genes or pathways in fibromyalgia patients. Finally, the potential therapeutic relevance of TLR-related signaling and low-dose naltrexone was inferred from the network findings and previous studies and was not directly evaluated in the present study. Further experimental and clinical studies are required to validate these findings.

## Conclusion

Currently, the available treatment modalities for fibromyalgia do not result in complete symptom relief in all patients. In this study, we used a network pharmacology approach and bioinformatic tools to identify potential hub genes and biological pathways associated with fibromyalgia. The Toll-like receptor signaling pathway was among the significantly enriched pathways identified in our analysis, and TNF and IL6 were identified as the top two hub genes based on network centrality. These findings provide potential molecular targets and pathways for further investigation in preclinical and clinical studies.

The findings also provide a rationale for further investigation of TLR-related mechanisms and the potential role of low-dose naltrexone in fibromyalgia. However, experimental studies and appropriately designed clinical studies are required to validate these hypotheses and determine their therapeutic relevance. The identified hub genes, particularly TNF and IL6, may also be investigated in future clinical studies to determine whether their circulating levels are associated with fibromyalgia severity or prognosis.

## Data Availability

All data used in the present study are publicly available from the databases and resources cited in the manuscript. GeneCards, DisGeNET, OMIM, and PharmGKB were initially screened for fibromyalgia-associated genes. GeneCards and DisGeNET were subsequently used for the analysis. Protein association data were obtained from STRING, and functional enrichment analysis was performed using ShinyGO. No patient-level, individually identifiable data, clinical records, or human samples were used.

https://www.genecards.org/

https://disgenet.com/

https://omim.org/

https://www.clinpgx.org/

https://string-db.org/

